# Quantifying Academic Risk Factors for Student Depression Using WHO Frameworks: Odds Ratios, SHAP Explainability, and Tipping Point Analysis

**DOI:** 10.64898/2026.08.09.26360019

**Authors:** Tanvir Ahmed, Md. Rashid Al Asif

## Abstract

Depression has become a serious concern for students worldwide. Aligned with the WHO Helping Adolescents Thrive (HAT) Guidelines and the Social Determinants of Health (SDoH) model, this study isolates five academically relevant factors—academic pressure, work/study hours, study satisfaction, sleep duration, and financial stress—from a dataset of 27,880 university students in India and quantifies their associations with depression. Unlike prior work that maximises classification accuracy, this study prioritises interpretability: logistic regression provides odds ratios (OR) with 95% confidence intervals, Random Forest (RF) and XGBoost rank predictors by feature importance, and SHAP (SHapley Additive exPlanations) values extend the analysis to individual-level risk explanation. Sleep Duration is encoded as an ordinal integer (0–3) preserving its natural order, and Spearman correlations are computed on the original 27,880-row dataset prior to any resampling. SMOTE oversampling was applied exclusively to the training set, and performance was evaluated on the original imbalanced test set (n = 5,576). Both ensemble models achieve approximately 77–78% accuracy and an AUC of 0.845, confirmed by 5-fold pipeline cross-validation (CV AUC ∼ 0.843). Academic pressure is the dominant risk factor (OR = 2.271; RF importance = 0.481; mean |SHAP| = 0.174), while study satisfaction (OR = 0.796) and sleep duration (OR = 0.835) are protective. Interaction analysis reveals how depression risk is amplified by low sleep, high financial stress, and extended study hours: under average conditions the 50% risk threshold is crossed at Academic Pressure ≈ 3.04, and under compounded adverse conditions this tipping point shifts to even lower pressure values. These findings provide data-driven thresholds aligned with WHO-endorsed modifiable determinants to support early detection and institutional counselling.

## I. Introduction

Student mental health has emerged as a critical issue globally, with approximately 1 in 7 adolescents affected by depression and anxiety [1]. At the university level, academic-related factors—high workload, insufficient sleep, low study satisfaction, and financial strain—are primary drivers of depressive symptoms [2]–[4]. These determinants align with the WHO Helping Adolescents Thrive (HAT) Guidelines [5], which identify stress management, school-based emotional regulation, and poverty as key modifiable targets for adolescent mental health interventions, and with the Social Determinants of Health (SDoH) model [6], which places education quality and income security as upstream structural determinants of health outcomes.

WHO states that depression and anxiety are the leading causes of mental illness and disability among adolescents worldwide [1]. Academic stressors contribute significantly to this burden. Yet most recent studies focus on maximising classification accuracy using ML, without grounding feature selection in public health frameworks or providing clinically interpretable outputs [7], [8].

This study addresses that gap by: (i) isolating five academically relevant factors aligned with WHO HAT and SDoH frameworks; (ii) quantifying independent factor effects using odds ratios [9] with 95% confidence intervals; (iii) ranking predictors by feature importance across two ensemble models; (iv) extending global explanations to individual-level risk profiles using SHAP values; and (v) identifying data-driven tipping points for early detection thresholds.

## II. Literature Review

Depression among university students has been reported across national and regional settings, with 29–44% attributing academic-related factors as primary contributors [10]–[12]. Academic pressure and high workload are consistently cited as major risk factors associated with burnout [13]–[15]. Sleep loss intensifies depressive symptoms [16], [17], study satisfaction serves as a protective factor [18], and financial pressure exacerbates vulnerability particularly in low-resource settings [19], [20].

### A. Limitations of existing ML approaches

Several studies have applied ML to predict student depression with high accuracy. Rois et al. [7] and Chowdhury et al. [21] achieved strong results with Bangladeshi student populations, while Zhai et al. [22] applied XGBoost and RF to a large US population (n = 61,619, AUC 0.74–0.77). Baba and Bunji [8] applied LR, RF, XGBoost, and LightGBM to Japanese university students with feature importance analysis. Most recently, Omarbekova et al. [23] proposed an XAI framework combining RF, LR, SHAP, and Friedman statistical testing for multi-class academic stress classification across two Kaggle datasets, achieving AUC up to 0.985. However, their study targets stress severity rather than depression, selects features statistically rather than through public health criteria, and reports neither odds ratios nor clinical tipping points (Table I).

**Table I.**
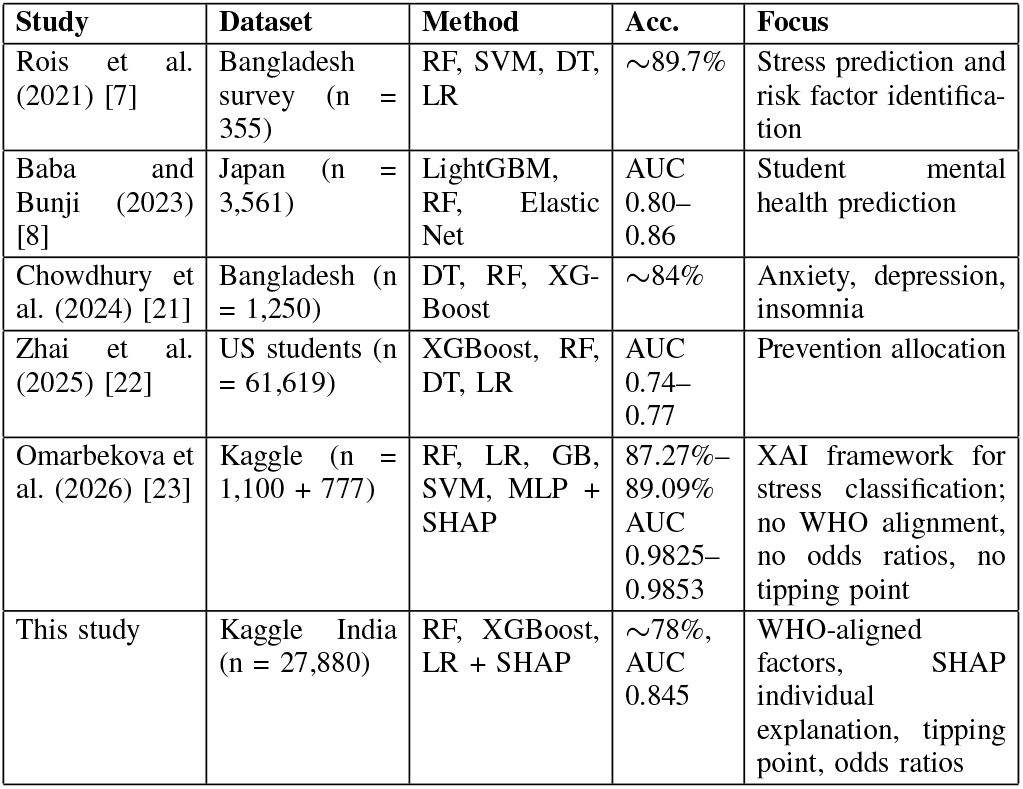
comparative analysis of related studies.

| Study | Dataset | Method | Acc. | Focus |
|---|---|---|---|---|
| Rois et al. (2021) [7] | Bangladesh survey (n = 355) | RF, SVM, DT, LR | ∼89.7% | Stress prediction and risk factor identifica-tion |
| Baba and Bunji (2023) [8] | Japan (n = 3,561) | LightGBM, RF, Elastic Net | AUC 0.80–0.86 | Student mental health prediction |
| Chowdhury et al. (2024) [21] | Bangladesh (n = 1,250) | DT, RF, XG-Boost | ∼84% | Anxiety, depression, insomnia |
| Zhai et al. (2025) [22] | US students (n = 61,619) | XGBoost, RF, DT, LR | AUC 0.74–0.77 | Prevention allocation |
| Omarbekova et al. (2026) [23] | Kaggle (n = 1,100 + 777) | RF, LR, GB, SVM, MLP + SHAP | 87.27%–89.09% AUC 0.9825–0.9853 | XAI framework for stress classification; no WHO alignment, no odds ratios, no tipping point |
| This study | Kaggle India (n = 27,880) | RF, XGBoost, LR + SHAP | ∼78%, AUC 0.845 | WHO-aligned factors, SHAP individual explanation, tipping point, odds ratios |

### B. WHO frameworks as a basis for factor selection

The WHO HAT Guidelines [5] provide evidence-informed recommendations on psychosocial interventions for adolescents. Recommendation A specifically advocates universally delivered interventions covering stress management, emotional regulation, and problem-solving in school settings directly implicating academic pressure and workload as modifiable targets. Recommendation D emphasises early indicated interventions for adolescents displaying emotional symptoms including depression. The HAT Guidelines also explicitly identify poverty as a priority adversity (Question 2b), linking financial stress to adolescent mental health vulnerability. The SDoH model [6], [24] places education quality and income/social protection as upstream structural health determinants. UNESCO’s Happy Schools framework [25], [26] further highlights workload balance and academic satisfaction as burnout prevention strategies. These frameworks collectively justify the five factors selected in this study.

### C. Research Contribution

No prior study has systematically (i) grounded feature selection in WHO HAT and SDoH criteria, (ii) quantified independent effects via odds ratios, (iii) ranked predictors across ensemble models, (iv) extended to SHAP individual-level profiles, and (v) identified clinical tipping points. This study addresses all five gaps.

## III. Methodology

This section explains the method applied in this research, investigating the relation between academic factors and depression under the framework of health organizations (Fig. 1).

**Fig. 1.**
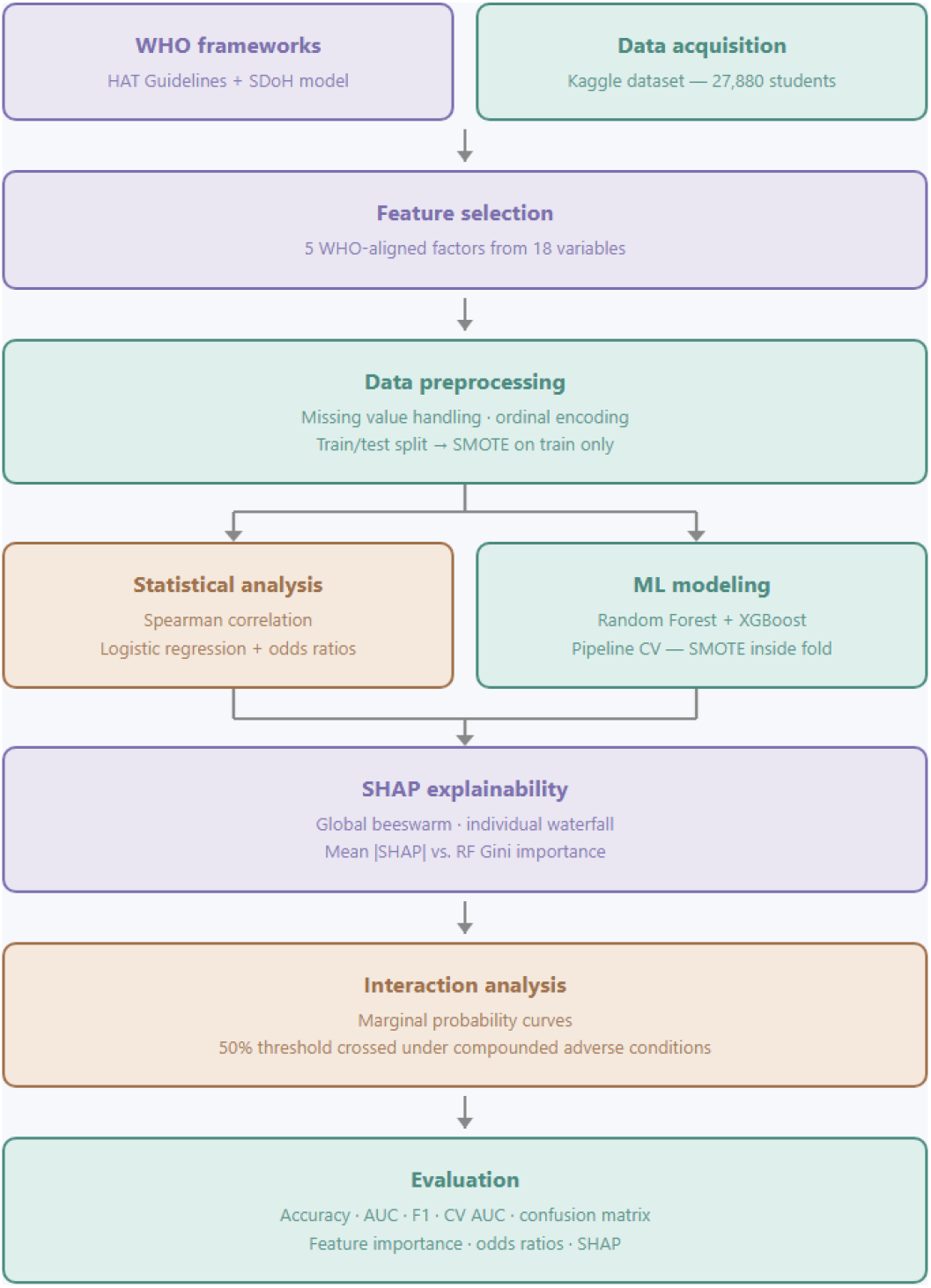
Overview of the proposed methodology. Starts with the data source and framework choice, then proceeds to data preprocessing. After that, statistical analysis, machine learning modeling, and interaction analysis are conducted, and the output is evaluated using interpretability and performance metrics.

### A. Data Source and Sampling

The “Student Depression Dataset” from Kaggle [27] contains demographic, academic, and lifestyle data for 27,883 Indian university students. Five factors were selected in alignment with WHO HAT and SDoH frameworks: Academic Pressure (1–5), Work/Study Hours (continuous), Study Satisfaction (1–5), Sleep Duration (ordinal integer: *<*5 h = 0, 5–6 h = 1, 7–8 h = 2, *>*8 h = 3), and Financial Stress (1–5). Depression is the binary target. After filtering and cleaning, 27,880 rows remain. The study relies on existing data, acknowledging self-report bias, consistent with WHO mental health surveillance guidelines.

### B. Data Preprocessing

Missing values were replaced with NaN and columns coerced to numeric types. Rows with missing values were dropped (3 rows removed). Sleep Duration was encoded as an ordinal integer (*<*5 h = 0, 5–6 h = 1, 7–8 h = 2, *>*8 h = 3), preserving its natural order and enabling a single, valid Spearman correlation coefficient to be reported; this is consistent with CDC and WHO sleep health recommendations [28]. To prevent data leakage, the dataset was first split into training (80%, n = 22,304) and test (20%, n = 5,576) sets using stratified sampling. SMOTE (random state = 42) was then applied exclusively to the training set, balancing it to 26,122 samples (13,061 per class). The test set retained the original imbalanced distribution (3,265 depressed, 2,311 non-depressed).

### C. Statistical Analysis

Spearman’s rank correlation was computed on the original 27,880-row cleaned dataset prior to any train/test split or SMOTE resampling, ensuring that the reported coefficients reflect the true population distribution rather than a synthetically balanced one. Logistic regression was fitted using statsmodels to estimate odds ratios with 95% confidence intervals, using maximum likelihood estimation. P-values were evaluated at *α* = 0.001, consistent with WHO’s emphasis on identifying modifiable risk factors for targeted intervention.

### D. Machine Learning Models

RF and XGBoost were selected for their complementary inductive biases: RF addresses variance through bagging, XGBoost addresses bias through sequential boosting. RF: n estimators = 500, max depth = 10, random state = 42. XGBoost: n estimators = 100, learning rate = 0.3, max depth = 5, random state = 42. Both models were trained on the SMOTE-balanced training set and evaluated on the original imbalanced test set.

### E. Cross-Validation

To avoid data leakage in cross-validation, an imblearn Pipeline was constructed embedding SMOTE within each fold of a 5-fold stratified cross-validation. SMOTE is fitted only on each fold’s training partition and never sees validation data. Reported metrics include mean accuracy, weighted F1-score, and AUC across folds.

### F. SHAP-Based Individual Risk Explanation

SHAP TreeExplainer was applied to the RF model on 500 held-out test instances. A global beeswarm plot illustrates each feature’s directional effect; a waterfall plot provides individual-level explanation for the highest-risk student (Academic Pressure = 5, Work/Study Hours = 11, Study Satisfaction = 1, Sleep Duration = 0 [i.e., *<*5 hrs], Financial Stress = 5), enabling counsellors to communicate personalised risk drivers.

### G. Interaction Analysis

Marginal probability curves were generated using a logistic regression model (LR) fitted on the SMOTE-balanced training set, varying Academic Pressure across 500 equally spaced values (1–5) while holding all other variables at their training-set means. Logistic regression was selected for interaction curves because its sigmoid output is mathematically continuous, producing smooth S-curves without discrete boundary artefacts. Interaction curves were generated by setting each secondary variable to its low and high extremes, revealing how sleep, satisfaction, financial stress, and study hours modulate depression probability across the full pressure range. The 50% threshold defines the tipping point for each condition.

## IV. Results

After cleaning, 27,880 rows remain (16,326 depressed, 11,554 non-depressed). After SMOTE on training data only, the training set contains 26,122 balanced samples; the test set retains the original imbalanced distribution (5,576 samples).

### A. Spearman Correlation

Table II presents Spearman correlation coefficients computed on the original 27,880-row dataset prior to resampling. Academic Pressure shows the strongest association (*ρ* = 0.472, moderate positive) and Financial Stress follows (*ρ* = 0.363). Work/Study Hours shows a weak positive correlation (*ρ* = 0.201), while Study Satisfaction (*ρ* = -0.168) and Sleep Duration (*ρ* = -0.087) show protective negative correlations. All correlations are significant at p *<* 0.001. Near-zero inter-feature correlations in the heatmap (Fig. 2) confirm low multicollinearity.

**TABLE II.**
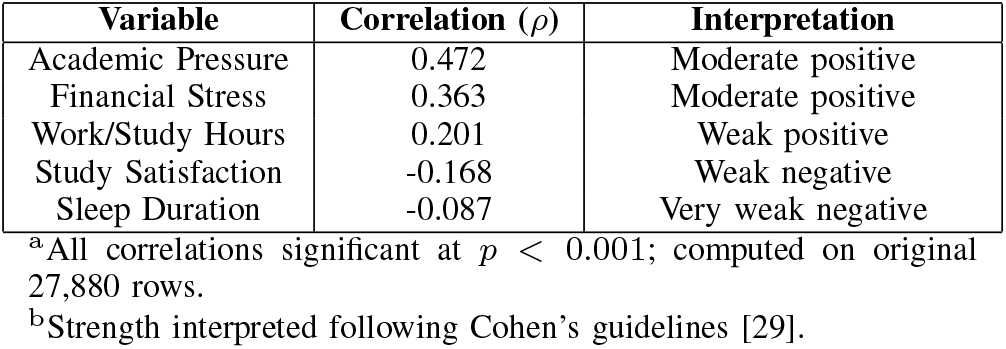
Spearman Correlation Coefficients with Depression (Original Data, Pre-SMOTE)

| **Variable** | **Correlation (*ρ*)** | **Interpretation** |
|---|---|---|
| Academic Pressure | 0.472 | Moderate positive |
| Financial Stress | 0.363 | Moderate positive |
| Work/Study Hours | 0.201 | Weak positive |
| Study Satisfaction | -0.168 | Weak negative |
| Sleep Duration | -0.087 | Very weak negative |

<sup>a</sup>All correlations significant at *p* < 0.001; computed on original 27,880 rows.
<sup>b</sup>Strength interpreted following Cohen's guidelines [29].

**Fig. 2.**
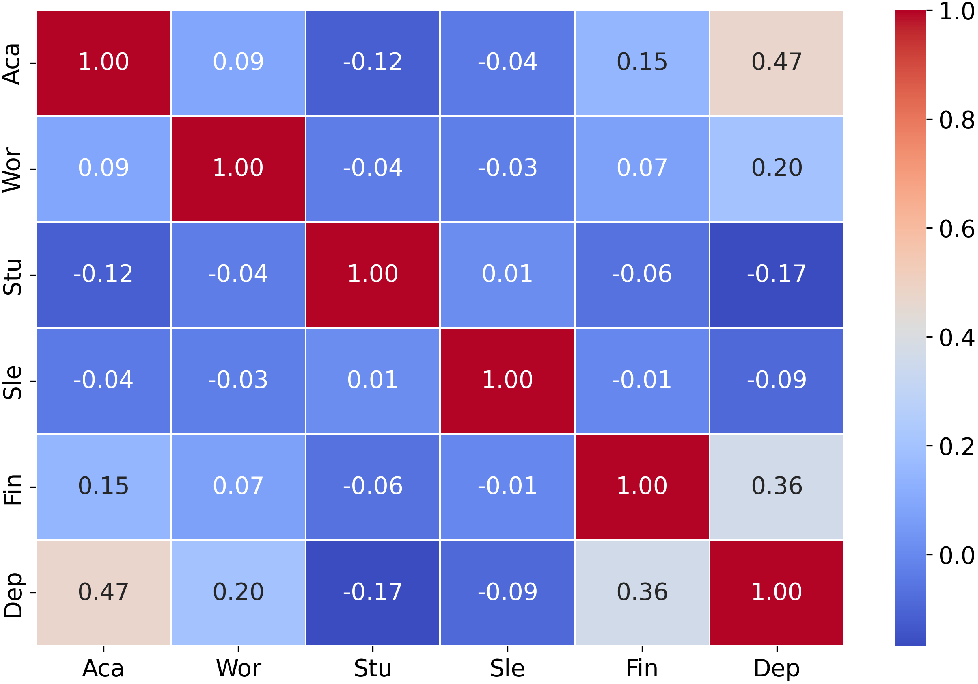
Spearman Correlation Matrix: Associations between academic factors (pressure, work/study hours, satisfaction, sleep duration), financial stress, and depression, computed on the original 27,880-row dataset prior to SMOTE resampling. Values range from ™1 (strong negative, blue) to +1 (strong positive, red). Abbreviations: Aca, Academic Pressure; Wor, Work/Study Hours; Stu, Study Satisfaction; Sle, Sleep Duration; Fin, Financial Stress; Dep, Depression. Generated using Seaborn and Matplotlib.

### B. Logistic Regression (Odds Ratios)

The logistic regression model achieved pseudo-*R*^2^ = 0.3015 and converged in 6 iterations. Table III shows Academic Pressure has the highest odds ratio (OR = 2.271, 95% CI [2.215, 2.328], *p <* 0.001): each one-unit increase more than doubles the odds of depression. Financial Stress is the second strongest risk factor (OR = 1.783), while Work/Study Hours shows a modest effect (OR = 1.128). Study Satisfaction (OR = 0.796) and Sleep Duration (OR = 0.835) demonstrate statistically significant protective effects. The odds ratio is calculated as OR_*i*_ = exp(*β*_*i*_), with 95% CI [exp(*β*_*i*_ *™* 1.96 SE(*β*_*i*_)), exp(*β*_*i*_ + 1.96 SE(*β*_*i*_))].

**TABLE III.** Logistic Regression Results (Odds Ratios)

| **Predictor** | **Coefficient** | **Odds Ratio (95% CI)** | **p-value** |
|---|---|---|---|
| Academic Pressure | 0.820 | 2.271 (2.215–2.328) | < 0.001 |
| Financial Stress | 0.578 | 1.783 (1.744–1.824) | < 0.001 |
| Work/Study Hours | 0.121 | 1.128 (1.119–1.138) | < 0.001 |
| Study Satisfaction | -0.229 | 0.796 (0.778–0.814) | < 0.001 |
| Sleep Duration | -0.180 | 0.835 (0.813–0.858) | < 0.001 |

<sup>a</sup>Model pseudo-R<sup>2</sup> = 0.3015, converged in 6 iterations.
<sup>b</sup>Sleep Duration encoded as ordinal integer (0–3); OR reflects a one-category-step increase (e.g., <5 h → 5–6 h).

### C. Machine Learning Performance

Table IV summarises performance on the original imbalanced test set and 5-fold pipeline CV. Both RF and XGBoost achieve approximately 77–78% accuracy and AUC = 0.845, demonstrating strong discriminative ability. CV AUC values of 0.843 (RF) and 0.842 (XGBoost), with low standard deviations (±0.63% and ±0.52%), confirm model stability. The confusion matrix shows 2,583 true negatives, 2,565 true positives, 683 false positives, and 700 false negatives, Fig 3 illustrates the ROC AUC curves. Class 1 (depressed) F1-scores of 0.80– 0.81 indicate the model successfully identifies the majority of at-risk students.

**TABLE IV.** Performance of Machine Learning Models.

| **Model** | **Test Acc.** | **Test AUC** | **CV Acc.** | **CV AUC** | **SD** |
|---|---|---|---|---|---|
| RF | 77.30% | 0.8450 | 77.21% | 0.8430 | ±0.63% |
| XGBoost | 77.64% | 0.8451 | 77.12% | 0.8416 | ±0.52% |

<sup>b</sup>Model performance on original imbalanced test set (n = 5,576) and 5-fold pipeline CV. SMOTE was applied only within training folds.

**Fig. 3.**
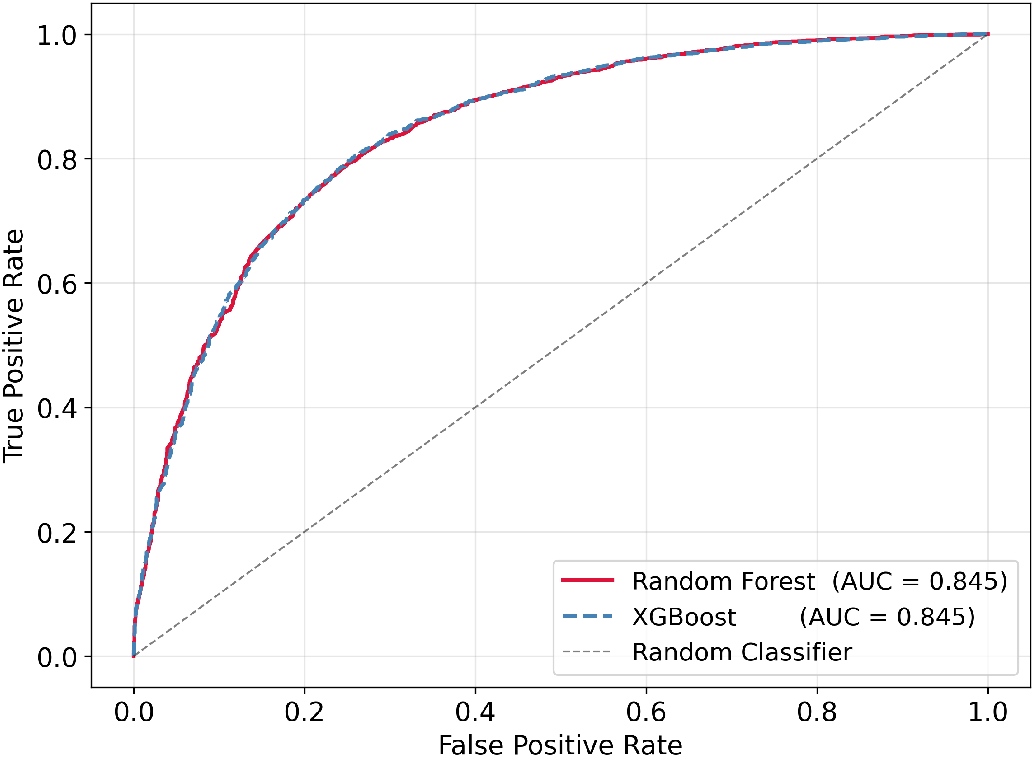
ROC curves for Random Forest and XGBoost on the original test set (n = 5,576), both achieving AUC = 0.845. Near-identical AUC values across two models confirm robustness of the WHO-aligned feature set. Generated using Matplotlib.

**Fig. 4.**
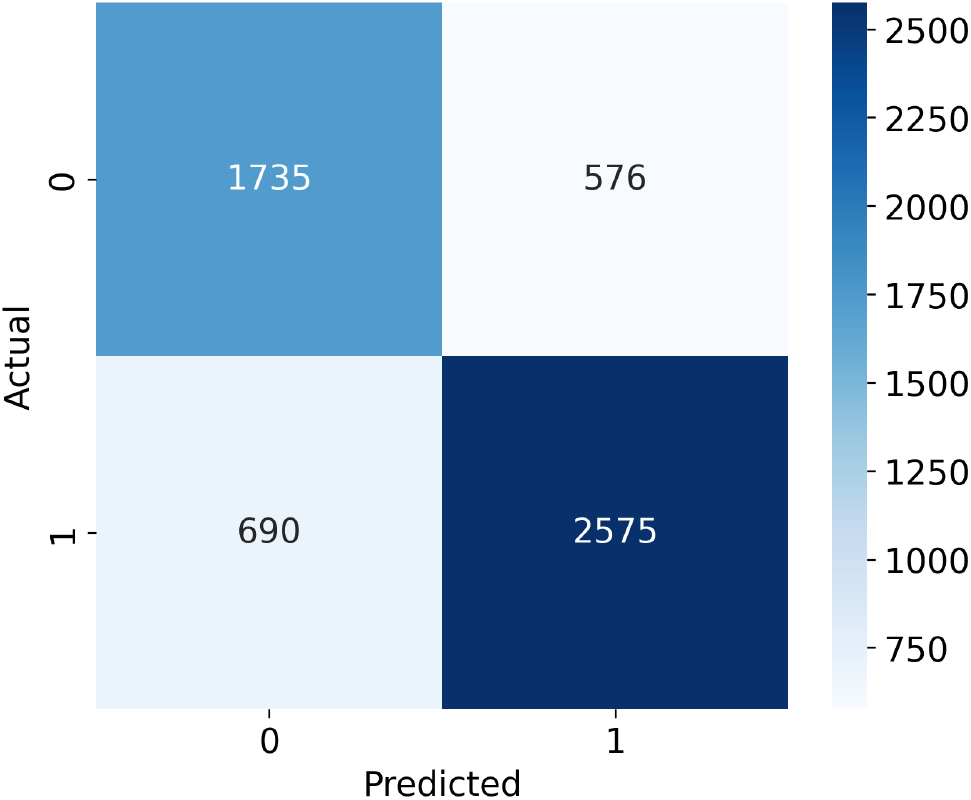
Confusion matrix for the Random Forest model on the original imbalanced test set (n = 5,576): 2,583 true negatives, 2,565 true positives, 683 false positives, and 700 false negatives. The higher recall for the depressed class (Class 1) reflects appropriate sensitivity for a mental health screening tool. Generated using Seaborn and Matplotlib.

### D. Feature Importance

Feature importance analysis consistently ranked Academic Pressure as the dominant predictor across both models (RF: 0.481; XGBoost: 0.500), followed by Financial Stress (RF: 0.273; XGBoost: 0.282). Work/Study Hours, Study Satisfaction, and Sleep Duration followed in descending order for RF; XGBoost assigned slightly higher importance to Sleep Duration over Work/Study Hours. Consistency across two models with different inductive biases strengthens the robustness of these rankings (Figs. 5–6).

**Fig. 5.**
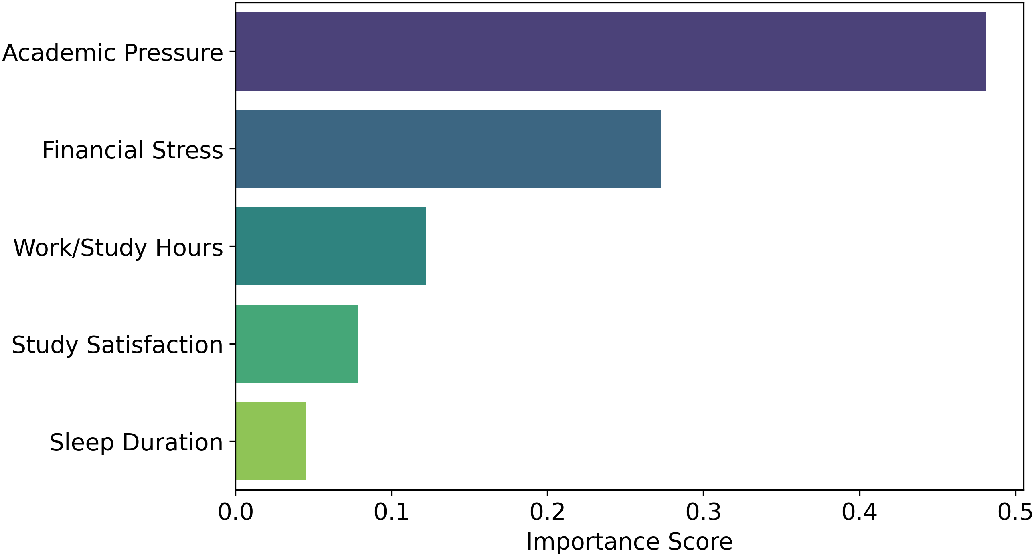
Feature importance scores from the RF model, ranked in descending order. Academic pressure emerges as the dominant predictor (0.481). Generated using Matplotlib.

**Fig. 6.**
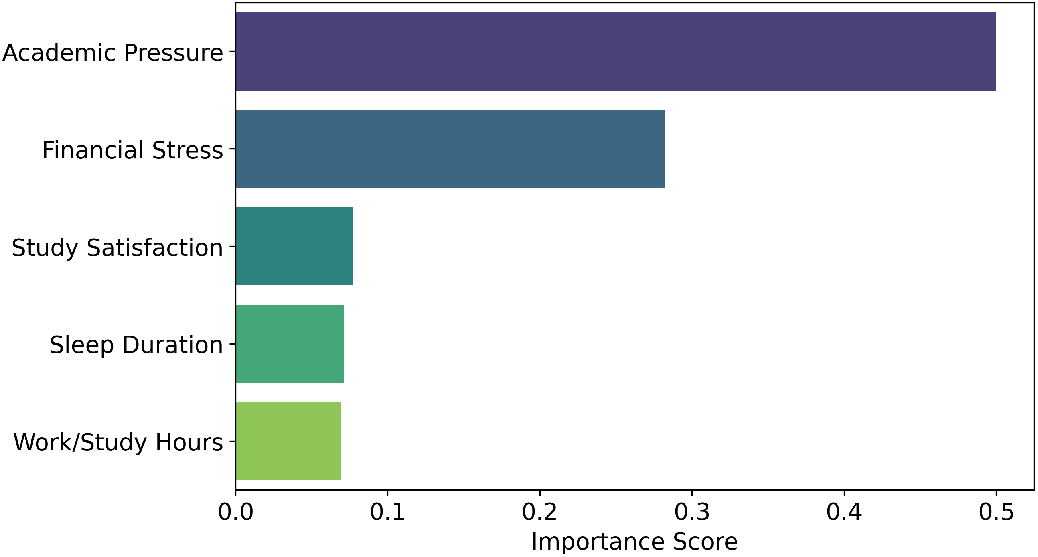
Feature importance scores from the XGBoost model, ranked in descending order. Academic pressure leads as the primary driver (0.500). Generated using Matplotlib.

### E. SHAP Explainability

Table V compares mean |SHAP| values with RF Gini importance. The rankings are consistent: Academic Pressure leads (mean |SHAP| = 0.1745), followed by Financial Stress (0.1409). The global beeswarm plot (Fig. 7) shows that high Academic Pressure values (red) consistently produce positive SHAP contributions pushing predictions toward depression, while low values (blue) suppress risk. Study Satisfaction and Sleep Duration show negative SHAP contributions for high values, confirming protective effects at the individual level—a directional nuance that Gini importance alone cannot capture.

**Table V.** global shap vs. rf gini feature importance.

| **Feature** | **Mean \|SHAP\|** | **RF Importance** |
|---|---|---|
| Academic Pressure | 0.1745 | 0.481 |
| Financial Stress | 0.1409 | 0.273 |
| Work/Study Hours | 0.0692 | 0.122 |
| Study Satisfaction | 0.0486 | 0.079 |
| Sleep Duration | 0.0258 | 0.046 |

**Fig. 7.**
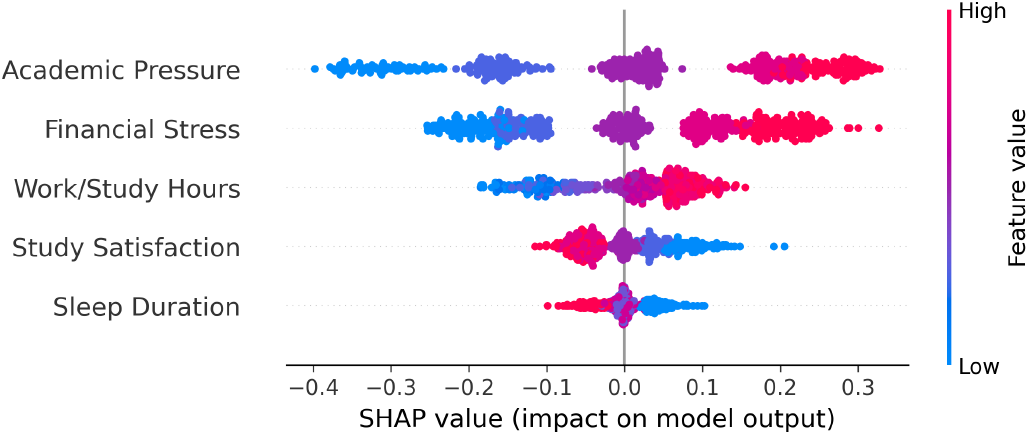
Global SHAP beeswarm plot for the RF model (500 test instances). Each point represents one student; horizontal position indicates feature contribution to the depression prediction; color indicates feature value (red = high, blue = low). Academic Pressure shows the largest mean absolute SHAP value with a consistent positive direction, while Study Satisfaction and Sleep Duration show negative contributions at high values, confirming their protective roles at the individual level. Generated using the SHAP library.

**Fig. 8.**
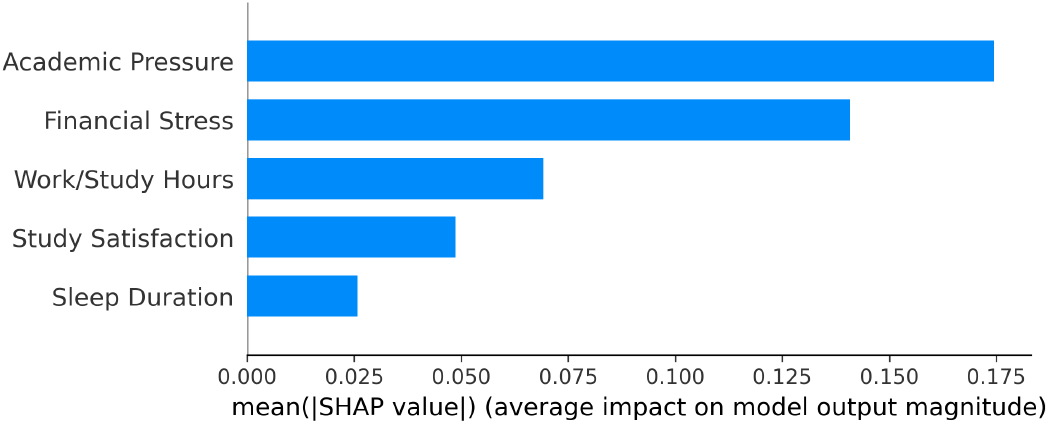
Global SHAP bar plot showing mean |SHAP| values per feature across 500 test instances. Academic Pressure (0.1745) and Financial Stress (0.1409) together account for over 70% of total explanatory weight, consistent with feature importance rankings from both RF and XGBoost. Generated using the SHAP library.

The waterfall plot (Fig. 9) for the highest-risk student (Academic Pressure = 5, Work/Study Hours = 11, Study Satisfaction = 1, Sleep *<* 5 hrs, Financial Stress = 5) shows all five features pushing the predicted probability substantially above the base rate. This individual-level explanation enables a counsellor to communicate to a specific student which factor contributes most to their personalised risk—a clinical utility not achievable through global importance scores alone.

**Fig. 9.**
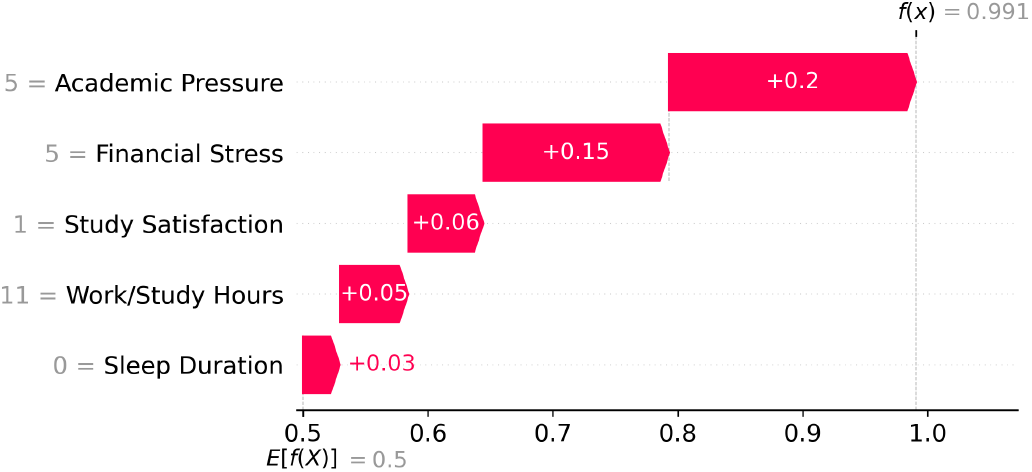
SHAP waterfall plot for the highest-risk student (Academic Pressure = 5, Work/Study Hours = 11, Study Satisfaction = 1, Sleep Duration = 0 [*<*5 hrs, ordinal scale], Financial Stress = 5). All five features push the predicted probability above the base rate, demonstrating how compounded adverse conditions produce maximum risk. Generated using the SHAP library.

### F. Tipping Point and Interaction Analysis

The marginal probability curve (Fig. 10), generated using a logistic regression model fitted on the SMOTE-balanced training set with 500 equally spaced pressure values, shows a monotonically increasing depression probability with academic pressure. When other variables are held at their training-set means, the curve crosses the 50% threshold at Academic Pressure ≈ 3.04—indicating that moderate academic pressure alone, even at average levels of all other risk factors, is sufficient to place a student in the majority-risk category. This tipping point of 3.04 on a 1–5 scale has direct institutional relevance: students reporting academic pressure at or above the midpoint of the scale already exceed the 50% estimated depression probability. Interaction analysis (Fig. 11–14) shows that under compounded adverse conditions (low sleep, high financial stress), the 50% threshold is crossed at even lower pressure values. Institutions can use pressure ≥ 3 as an early-warning criterion for proactive counselling outreach, particularly for students also reporting financial hardship or poor sleep.

**Fig. 10.**
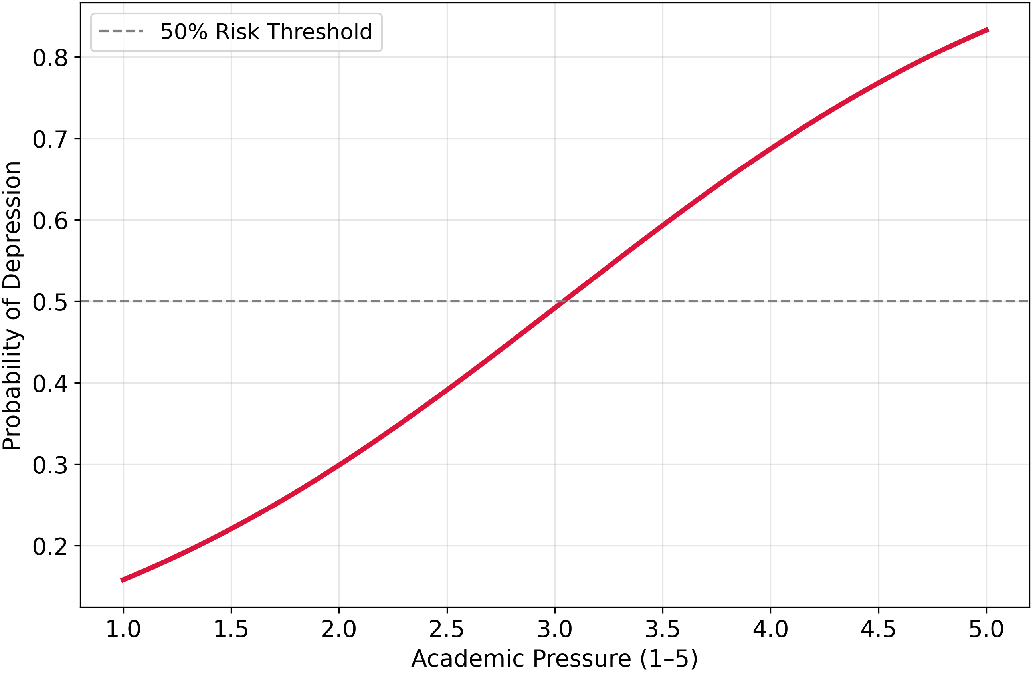
Marginal probability curve illustrating depression risk as Academic Pressure increases (scale 1–5), with all other variables fixed at training-set means. Generated from a logistic regression model fitted on the SMOTE-balanced training set using 500 equally spaced pressure values, producing a smooth sigmoid curve. Under average conditions the curve crosses the 50% risk threshold at Academic Pressure ≈ 3.04; compounded adverse conditions (see interaction plots) shift the threshold lower. Generated using Matplotlib and scikit-learn.

**Fig. 11.**
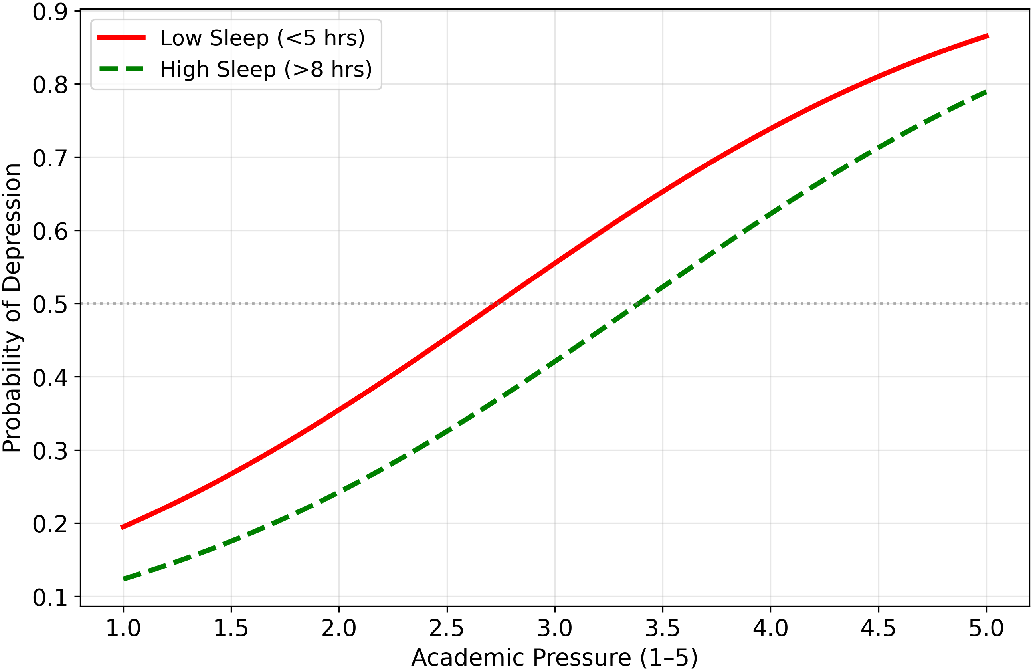
Interaction plot: Effect of Sleep Duration on depression probability across Academic Pressure levels. Low sleep (*<*5 hrs, ordinal = 0) substantially elevates depression risk at all pressure levels compared to high sleep (*>*8 hrs, ordinal = 3). Generated using Matplotlib.

**Fig. 12.**
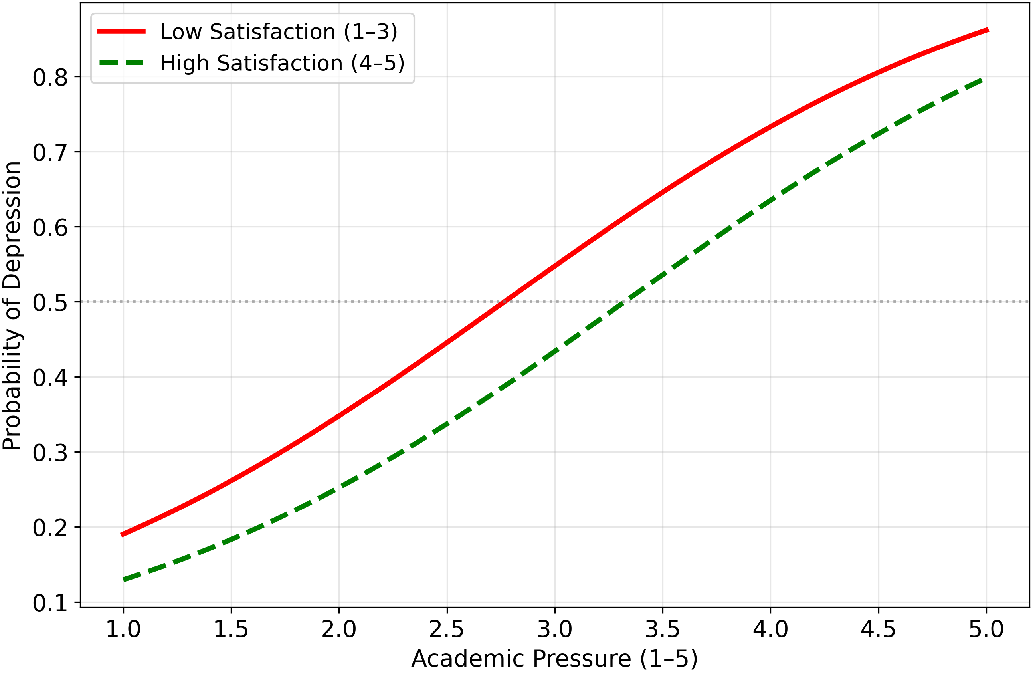
Interaction plot: Effect of Study Satisfaction on depression probability across Academic Pressure levels. Low satisfaction (score 1–3) consistently elevates risk by approximately 10–15 percentage points over high satisfaction (4–5) at every pressure level. Generated using Matplotlib.

**Fig. 13.**
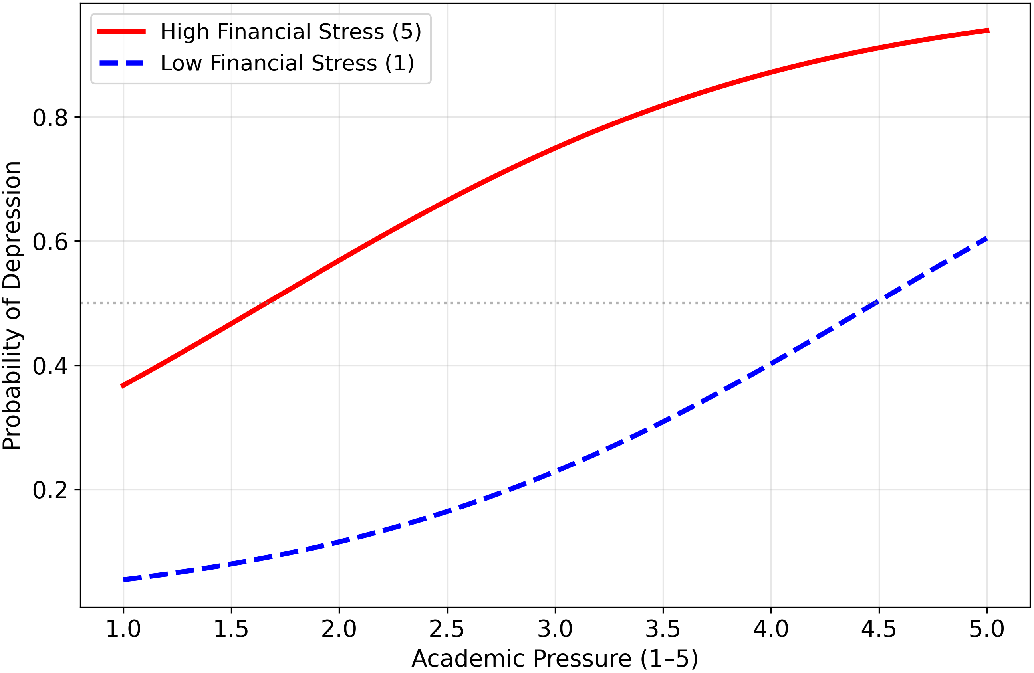
Interaction plot: Effect of Financial Stress on depression probability across Academic Pressure levels. High financial stress (score 5) crosses the 50% threshold at substantially lower academic pressure than low financial stress (score 1), consistent with the WHO SDoH model’s framing of income insecurity as an upstream health determinant. Generated using Matplotlib.

**Fig. 14.**
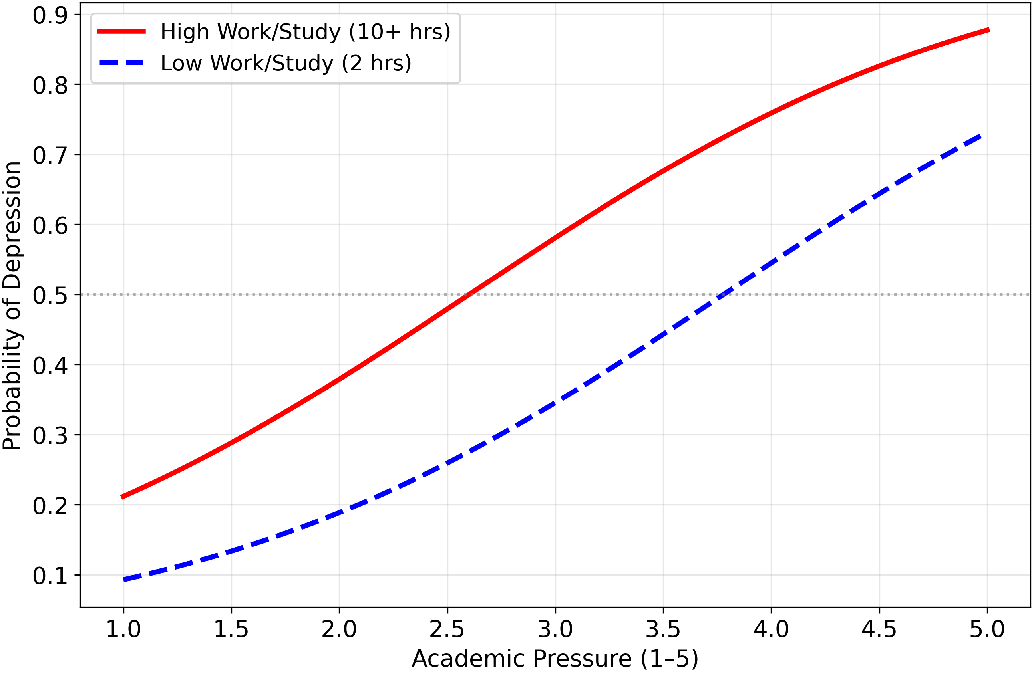
Interaction plot: Effect of Work/Study Hours on depression probability across Academic Pressure levels. Extended hours (*>*10 hrs) shift the entire probability curve upward relative to low hours (2 hrs), consistent with HAT Recommendation A’s emphasis on workload balance as a modifiable determinant. Generated using Matplotlib.

Interaction analysis reveals how contextual factors modulate depression risk (Figs. 11–14). Students with low sleep (*<* 5 hours, ordinal 0) exhibit substantially elevated probability curves across all pressure levels compared to high-sleep students (*>* 8 hours, ordinal 3), with probabilities approaching 0.65–0.70 at moderate pressure, far exceeding the 50% threshold earlier than the mean condition. Low study satisfaction (1–3) consistently elevates depression probability by approximately 10–15 percentage points over high satisfaction (4–5) at every pressure level. High financial stress (5) shifts the 50% crossing point to substantially lower academic pressure values compared to low financial stress (1), consistent with the SDoH model’s framing of income insecurity as an upstream determinant. Extended work/study hours (10+ hours) shift the entire probability curve upward relative to low hours (2 hours), evidencing cumulative burnout consistent with HAT Recommendation A’s emphasis on workload balance.

## V. Discussion

The results confirm that five WHO-aligned academic factors collectively explain a substantial portion of student depression risk (pseudo-*R*^2^ = 0.3015, AUC = 0.845). The marginal probability analysis reveals that academic pressure alone, even at average levels of other variables, crosses the 50% depression threshold at a pressure score of approximately 3.04—underscoring that depression risk is not confined to extreme academic stress but emerges at moderate levels for the average student. This is a substantive finding: because 3.04 falls just above the midpoint of the 1–5 scale, any student reporting academic pressure at or above the neutral midpoint is already estimated to be in the majority-risk category under average conditions. Interaction analysis further demonstrates that low sleep or high financial stress lower this tipping point further, providing institutions with a concrete, multifactor screening criterion consistent with the HAT Guidelines’ Recommendation D emphasis on early indicated interventions for adolescents with emotional symptoms.

Financial Stress emerges as a secondary but substantial pathway, operating through the SDoH model’s structural mechanism. The amplification effect in the interaction curves, high financial stress dramatically lowers the pressure threshold at which students enter the *>* 50% risk zone, highlights that socioeconomic interventions (bursaries, emergency funds, financial counselling) are complementary to, not substitutes for, academic workload management.

The protective roles of study satisfaction (OR = 0.796) and sleep duration (OR = 0.835) align with HAT Recommendation A’s emphasis on school environments that foster positive engagement. Interaction plots demonstrate that these factors widen the safe operating zone for academic pressure, suggesting that satisfaction and sleep hygiene interventions could meaningfully delay onset of high-risk states.

The SHAP analysis adds clinical utility absent in previous studies. While global feature importance identifies average predictor influence, the waterfall plot demonstrates that a counsellor can explain to an individual student precisely how each aspect of their academic profile contributes to their depression risk. This personalised explanation is directly applicable in university mental health settings.

Compared to prior work achieving 74–89% accuracy [7], [8], [21], [22], the present study’s 77–78% accuracy on a genuinely imbalanced test set, with SMOTE applied only to training data, is an honest and robust estimate. The AUC of 0.845, achieved using only five theoretically grounded features rather than all survey variables, demonstrates that WHO-aligned feature selection does not compromise predictive power while substantially improving interpretability and policy relevance.

Limitations include self-reported data (response bias risk), cross-sectional design precluding causal inference, and a single-country dataset. The binary depression label is not derived from a validated clinical scale such as PHQ-9, which constrains clinical interpretability. Future work should incorporate validated psychometric instruments, multi-country datasets, and longitudinal designs. Mediation analysis could clarify whether academic pressure operates directly or through intermediate variables such as sleep deprivation, per WHO’s mhGAP guidelines [30].

## VI. CONCLUSION

This paper quantifies five WHO-aligned academic risk factors for student depression using logistic regression, Random Forest, XGBoost, and SHAP explainability. Spearman correlations computed on the original 27,880-row dataset and ordinal encoding of Sleep Duration ensure methodological rigour. The key finding—that depression probability rises monotonically with academic pressure and crosses the 50% threshold at Academic Pressure ≈ 3.04 under average conditions, and at even lower values under compounded adverse conditions (low sleep or high financial stress)—provides institutions and counsellors with actionable, data-driven screening criteria grounded in WHO HAT Guidelines and the SDoH model. Financial stress, low study satisfaction, insufficient sleep, and extended study hours compound this risk through distinct but interacting mechanisms, each quantified through SHAP contributions and interaction probability curves. The extension of global feature importance to individual-level SHAP explanations represents this study’s primary contribution over prior ML-based work. Future research should validate these findings longitudinally across multiple cultural contexts and explore integration of personalised SHAP-based risk profiles into university mental health monitoring systems.

## Data Availability

All data used in this study are publicly available online at: https://www.kaggle.com/datasets/hopesb/student-depression-dataset. All analysis code is available upon reasonable request to the authors.

